# Tune In or Take the Stage? A Randomized Controlled Trial Comparing After-School Music and Theatre Training with Neuroimaging Outcomes for Youth

**DOI:** 10.64898/2026.06.03.26354844

**Authors:** Kevin Jamey, Ellen Herschel, Caitlin Noel, Jed Villanueva, Melissa Reyes, Eustace Hsu, Beatriz Ilari, Wendy Mack, Shan Luo, Assal Habibi

## Abstract

**Introduction:** While growing evidence suggests that music training supports child development, few long-term randomized controlled trials (RCTs) have rigorously tested these claims. Moreover, it remains unclear whether the benefits are confined to music-specific domains or extend to higher-order cognitive functions such as inhibitory control (IC), a core executive function associated with long-term outcomes in academic achievement, career success, socio-emotional health, and physical well-being. This paper presents the protocol for the Extracurricular Activity and Child Early Learning and Development (EXCEL) trial, an RCT designed to assess the feasibility of a long-term music training program focusing on the brain and behavioral correlates of IC.

**Methods:** A total of 126 children, aged 6 to 8 years and residing in neighborhoods with limited resources in Los Angeles, were individually randomized to either a music (intervention) or theatre (active control) after-school program. Both programs were delivered over 24 months by established community arts organizations. Eligibility criteria included: average intellectual functioning, no major medical or psychiatric conditions, and MRI eligibility. Children with prior formal music training exceeding six months or severe hearing impairment were excluded. Before the intervention began, all participants completed baseline behavioral and neuroimaging assessments. The primary trial aim was to assess the effects of extended music training, relative to theatre training, on changes in measures of IC (i.e., Go/No-Go task and delayed gratification) and related neural functional activation. A secondary interim aim of the trial was to evaluate the feasibility of conducting a long-term RCT of music education in a first cohort, measured by participant retention, adherence to the program, willingness to continue at the 12-month mark, and fidelity.

**Progress:** Recruitment, screening, baseline testing, randomization, and program enrollment began August 1^st^, 2022, and after-school programming began October 1^st^, 2022. The randomized interventions and all data for the first cohort (N = 42) have been collected. Intervention and active control programs for a second cohort are ongoing and will end in Fall 2026.

**Discussion:** This paper reports the EXCEL trial protocol and provides feasibility estimates of implementing a long-term randomized controlled trial of music training in real-world, community-based settings with children. While similar neuroimaging RCTs are currently underway in Europe, the EXCEL trial is among the first in the United States to integrate longitudinal neuroimaging with arts intervention. Findings will inform the viability of scaling such programs and contribute to our understanding of how sustained music engagement may influence the development of inhibitory control circuitry in childhood.

The EXCEL Trial **Registration**: ClinicalTrials.gov NCT05502939

Protocol Version 1, April 22, 2026

## Music Training and Brain Plasticity

Formal music training has been linked to benefits for brain development. Studies show structural and functional differences between trained musicians and non-musicians in auditory, motor, and executive networks (Dalla Bella, 2016; Herholz & Zatorre, 2012; Zuk et al., 2014). Longitudinally, a randomized controlled trial (RCT) in 5- to 7-year-olds found that 15 months of training increased gray matter in motor regions (e.g., right precentral gyrus), auditory cortices (e.g., right Heschl’s gyrus), and the corpus callosum, with gains tied to musical and motor improvements (Hyde et al., 2009). Likewise, a non-randomized study of 6- to 9-year-olds showed that two years of training, compared to sports, strengthened frontal activation (e.g., inferior frontal gyrus) during executive tasks and optimized interhemispheric and temporal lobe development (Habibi, Damasio, Ilari, Sachs, et al., 2018; Habibi, Damasio, Ilari, Veiga, et al., 2018). These findings suggest music-induced brain plastic changes, however, replication through robust RCTs with sustained training and participant-level randomization is needed to confirm causality and guide education policy.

## Inhibitory Control as a Target of Music Training

Inhibitory control (IC) is an executive function often recruited during music practice and well suited for testing training effects induced by music-based activities. IC includes response inhibition and delayed gratification. Strong IC predicts academic achievement and socioemotional adjustment (Allan et al., 2014; Diamond, 2013; Diamond & Lee, 2011; Fogel et al., 2019; Fosco et al., 2019). Music training directly engages both IC components. Response inhibition is required to suppress errors, align motor actions with auditory feedback, and sustain attention across competing demands (Vuust et al., 2022; Zuk et al., 2014). Delayed gratification is embedded in the long-term structure of practice, where progress emerges only after sustained effort (Gebauer et al., 2012; Lehmann & Ericsson, 1997). Musicians themselves report that training fosters discipline, patience, and persistence (Campbell et al., 2007). These features actively recruit prefrontal and cingulo-opercular networks supporting IC (Garon et al., 2012; Niendam et al., 2012). Middle childhood (ages 6–12) is a sensitive period, as motor and perceptual skills for music emerge while the prefrontal cortex undergoes rapid maturation (Luna et al., 2010; Penhune, 2011).

## Behavioral Evidence Linking Music and IC

Behavioral findings support the role of music in strengthening IC. A meta-analysis of 22 studies found small-to-moderate effects of training on IC in children (Hedges’ g = 0.31), stronger in RCTs (g = 0.60; Jamey et al., 2024). Long-term community programs also enhance delayed gratification: children enrolled for 3–4 years showed greater ability to delay smaller rewards for larger future rewards than controls (Hennessy et al., 2019). These results suggest music training strengthens both response inhibition and reward-related self-control.

## Neuroimaging Evidence

IC relies on several brain regions, including the dorsolateral prefrontal cortex (dlPFC), anterior cingulate cortex (ACC), and supplementary/pre-supplementary motor areas (SMA/pre-SMA; Niendam et al., 2012; Swick et al., 2011). These regions mature substantially in childhood, supporting the regulation of impulses in pursuit of long-term goals (Luna et al., 2010). Other structured activities also enhance IC and modulate cognitive-control networks (Chen et al., 2014; Davis et al., 2011; Flook et al., 2010; Lakes & Hoyt, 2004; Voss et al., 2011)

Music training has been associated with similar neurodevelopmental effects. In a non-randomized longitudinal study, two years of group-based training were linked to changes in cortical maturation and increased white matter integrity, with functional differences observed in the IFG during inhibition tasks (Habibi, Damasio, Ilari, Veiga, et al., 2018). Even short interventions show effects: 20 days of computerized music training enhanced the N2 component in pre-school children, a marker of conflict monitoring, during an inhibition control task (Moreno et al., 2011). Cross-sectional fMRI studies show that trained musicians recruit fronto-parietal control networks more strongly during IC tasks (Zuk et al., 2014). In contrast, expert musicians display increased prefrontal–premotor connectivity during improvisation, suggesting refined top-down motor control (Pinho et al., 2014). Together, findings converge on the claim that music training influences IC-related circuitry. However, no long-term participant-level RCT with active controls and neuroimaging has yet been conducted in naturalistic settings, such as after-school programs.

## Gaps and Study Rationale

Without causal evidence, observed executive function changes may reflect pre-existing traits rather than effects of music training, underscoring the need for rigorous long-term randomized trials. Children from neighborhoods with limited resources remain largely excluded from such research (Noble et al., 2012).

The present protocol addresses this gap through a two-year parallel-arm RCT comparing after-school music and theatre training delivered by community arts organizations to children aged 6–8 from Los Angeles neighborhoods with limited resources. Beyond testing effects on IC and brain circuitry, the study will generate effect-size estimates for future trials, evaluate the feasibility of community-based RCTs, and address gaps in prior work that lacked participant-level randomization (Habibi et al., 2014, 2022).

## Materials and Methods

### Research Aims

We detail here the protocol for the Extracurricular Activity and Child Early Learning and Development (EXCEL) trial, a music RCT in children from communities with limited resources. The EXCEL trial feasibility measures obtained from a first trial cohort are summarized in this paper using:

1. Retention rate: the proportion of all randomized and enrolled participants in each group remaining active in the EXCEL trial after one year and at the end of the 2-year EXCEL trial, with a target of ≥70%.
2. Adherence: the proportion of attended sessions out of total planned sessions over one year, with a goal of ≥70% adherence per participant.
3. Commitment to continue: the number of students committed to continue for the second year of intervention/the total number of students randomized.
4. Fidelity: reports by instructors and external observers on classes, including interruptions, activities, engagement, and group work composition.

Feasibility and fidelity outcomes are assessed at 12 months and at the end of the 2-year EXCEL trial.

The overall aim of the EXCEL trial is to evaluate the impact of two years of music intervention, compared to an active control (theatre intervention), on behavioral and neural indices of IC. Key brain regions of interest (ROIs) consistently associated with IC activity include the dlPFC, ACC, and SMA/pre-SMA areas (Niendam et al., 2012; Swick et al., 2011). A two-year training period was selected based on evidence that behavioral improvements in IC typically emerge after extended durations of music training (e.g., Hennessy et al., 2019; Holochwost et al., 2017; Jaschke et al., 2018). This aligns with the proposal that music-related neuroplasticity may initially manifest in auditory and motor systems, with more distal transfer to higher-order cognitive regions requiring prolonged musical engagement.

We hypothesize that:

1. The music intervention will show greater improvements in IC than the theatre intervention, as measured by behavioral assessments.
2. Children receiving the music intervention will show greater functional activation of prefrontal IC regions during fMRI IC tasks compared to those receiving the theater intervention.
3. Behavioral changes in IC specific to those in the music intervention will correlate with functional neural activation changes in IC.
4. Secondary and exploratory hypotheses will investigate the effect of the music intervention on broader IC skills, delayed gratification, auditory processing, various musical abilities (rhythmic and melodic), academic achievement, and physical well-being. Additionally, changes in brain structure (cortical thickness, grey and white matter volumes) and connectivity (diffusion tensor imaging) will be examined.

### The EXCEL Trial Design

This project is funded by a National Institutes of Health (NIH) R61/R33 phased innovation grant. The R61 phase established feasibility, and the R33 phase evaluates efficacy. The EXCEL trial is a two-cohort, parallel-arm randomized controlled trial (RCT) conducted in community settings across Los Angeles, California. Cohort 1 (R61) recruited and randomized 42 children (21 per arm) between August 1^st^ and November 30^th^ 2022, with follow-up completed in December 2024. Cohort 2 (R33) began enrollment June 1^st^ 2024 and continued through March 30^th^ 2025, with a target of 84 participants (42 per arm). Figure 1 presents the SPIRIT schedule of enrollment, allocation, intervention delivery, and outcome assessments across the 24-month EXCEL trial.

**Figure 1.**
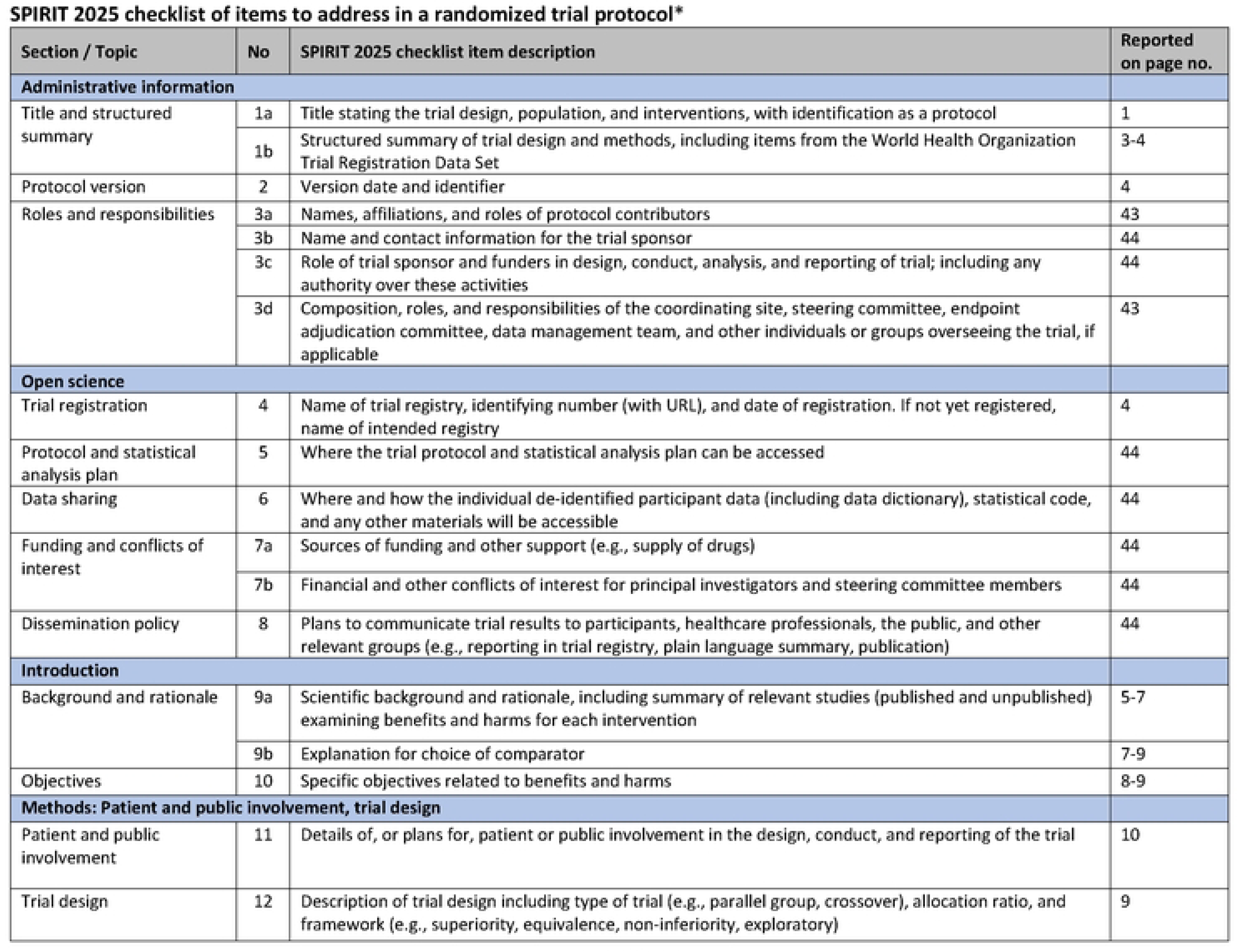
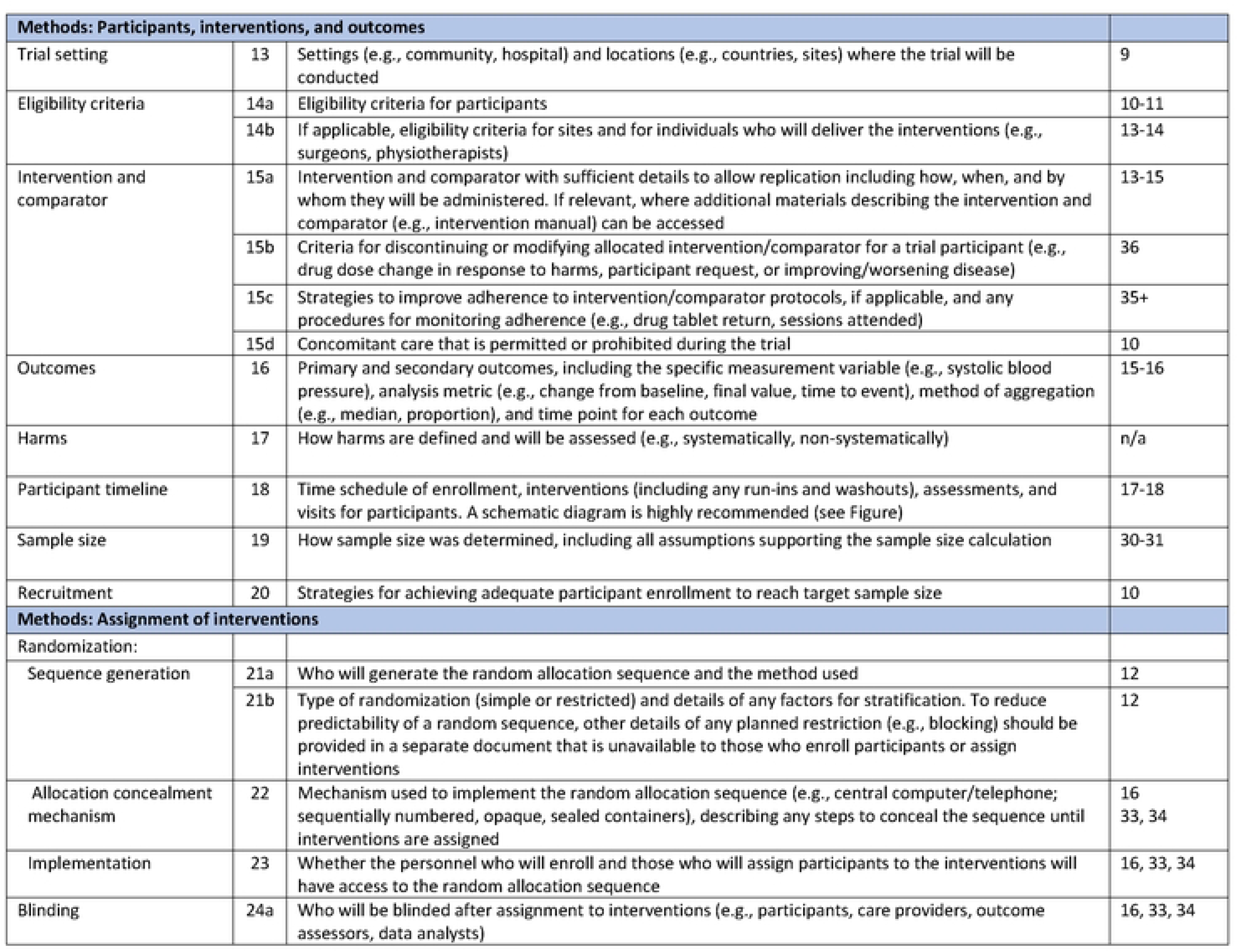
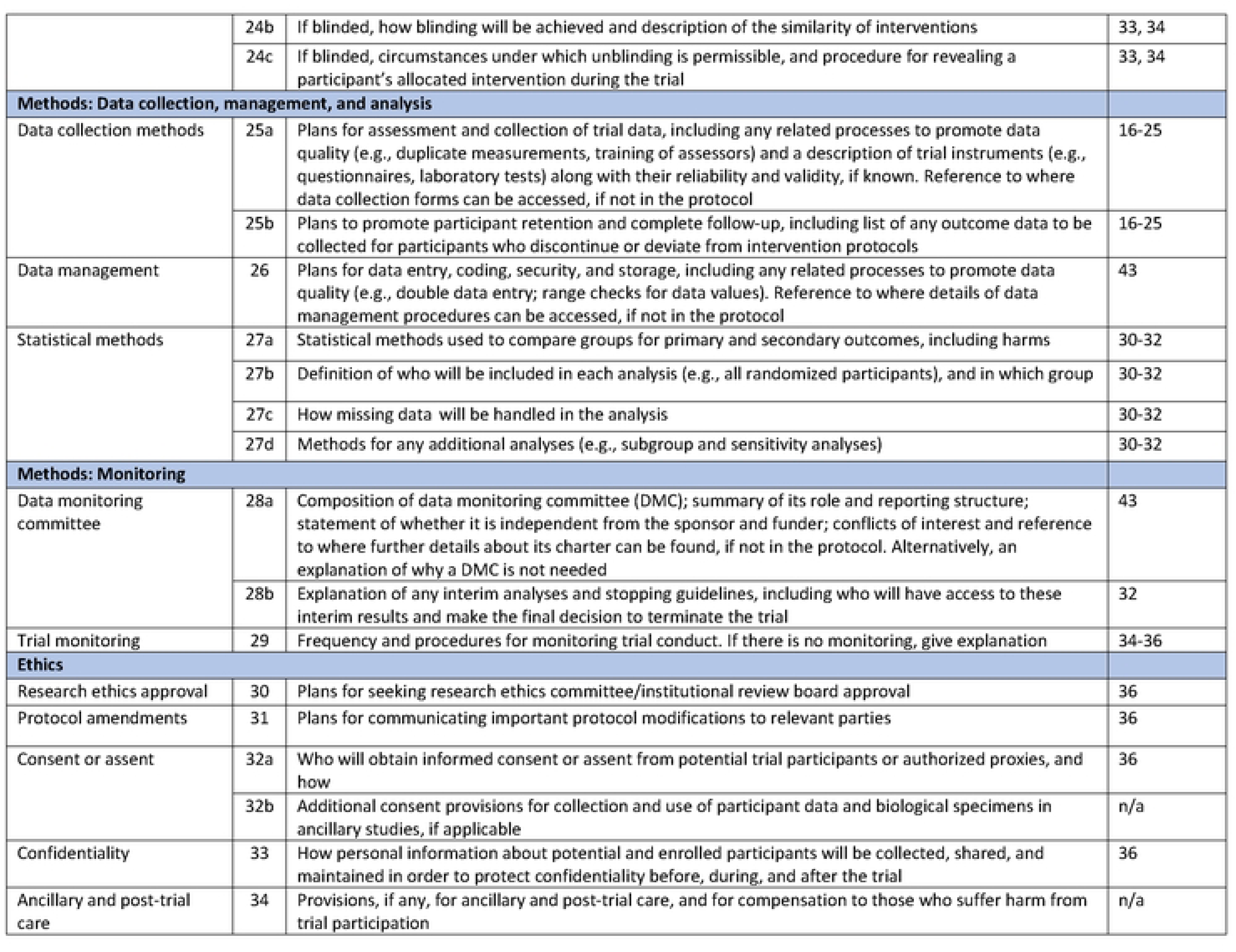
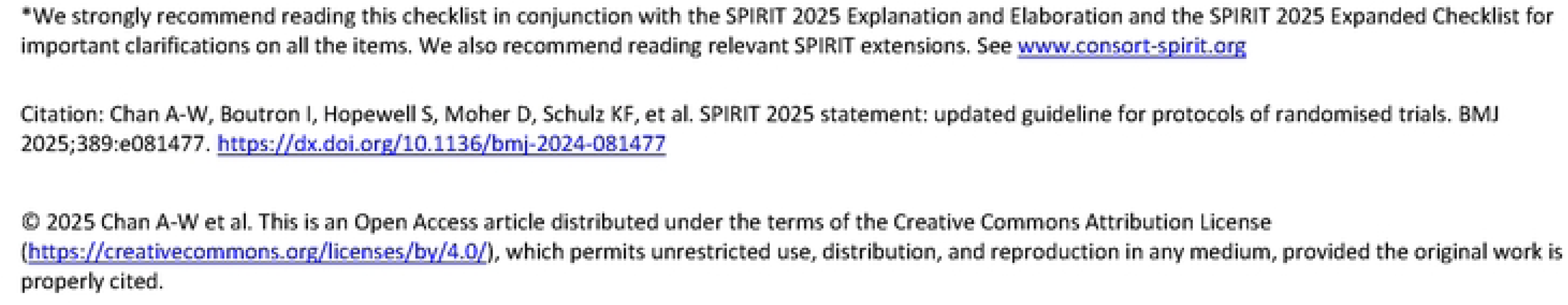
SPIRIT schedule of enrollment, interventions, and assessments for the EXCEL trial. Participants are screened and randomized to either the music or theatre intervention. Behavioral, questionnaire, and neuroimaging assessments are conducted according to the schedule shown at baseline, 6 months, 12 months, 18 months, and 24 months.

Participants are children aged 6–8 years from families with limited resources, defined as those earning below the median household income and residing in Los Angeles zip codes with limited resources (Kirkeby, 2021). The intervention arm receives a 24-month music-based after-school program. The active control arm receives a theatre-based program matched in duration and intensity. Assessments include behavioral and neuroimaging measures at baseline and 24 months, with additional behavioral testing at 12 months and questionnaires at 6 and 18 months. Research staff are blinded to group assignment, except for a limited number of coordinators handling family communication and logistics. The EXCEL trial is registered at ClinicalTrials.gov NCT05502939. Feasibility and fidelity metrics for each cohort are reported in this paper. Following program completion, training efficacy will be examined using musical and executive functioning outcome measures, but this is beyond the scope of this publication.

### Participant Recruitment and Screening

The EXCEL trial participants are children aged 6 to 8 years living within a 5-mile radius of the University of Southern California (USC) park campus, with no more than 6 months of prior formal music experience. Recruitment was conducted through social media (Facebook, Instagram), community outreach via local organizations and centers, flyer distribution to elementary schools, door-to-door outreach, and word-of-mouth referrals. The research team conducted eligibility screening via Zoom or in person. Children were eligible for inclusion if they met the following criteria:

1. Typical cognitive functioning (standard score > 85 on the Abbreviated Wechsler’s Scale of Intelligence, WASI-II)
2. Lived in a household with limited resources, defined as:

- (a) family income below the median annual income for their family size (Kirkeby, 2021); and
- (b) residence in a zip code where the average annual household income falls below the Los Angeles median for a family of four (> 68,000 USD in 2021)
3. No history of significant medical conditions
4. MRI eligible, as determined by standard psychological and physical safety criteria (e.g., absence of claustrophobia, metallic implants, pacemakers)
5. Between 6 and 8 years of age at the time of enrollment

Exclusion criteria were:

1. Current diagnosis of a neurological or psychiatric disorder, unless ADHD without regular medication intake
2. Previous formal music training exceeding six months
3. Untreated severe hearing or vision impairment
4. MRI ineligibility due to safety concerns (e.g., claustrophobia, metal implants, or incompatible dental work)

The EXCEL trial was designed to focus on households with limited resources and was representative of the ethnicity of the neighborhoods surrounding USC. A limited number of exceptions were approved by the EXCEL trial PI. Specifically, 22 participants (cohort 1: 10; cohort 2: 12) who lived outside the designated zip code radius were included, as their zip codes still met the income eligibility criteria. The furthest participant in cohort 1 lived 7 miles outside the radius; for cohort 2, the furthest was 20 miles away. See the CONSORT diagram (Figure 3) for details on the recruitment flow. No participant or public involvement in the design, conduct, and reporting of the trial is planned.

### Randomization

Participants who completed the screening assessments and met the inclusion and exclusion criteria were randomized before initiating interventions. Participants were randomized into a music or theatre group in a 1:1 allocation. Randomization was stratified on gender and age (<7, ≥7). To preserve the integrity of randomization and masking procedures, access to the randomization database was restricted to six staff members (one principal investigator, one collaborating professor, and two project coordinators per cohort). These individuals were not involved in any aspect of data collection or analysis.

### Screening Procedures

Participant screening is conducted in three stages. First, parents complete an interest form, which includes questions about the child’s age, socioeconomic background, and zip code. Second, families are contacted by the research team for a structured phone interview to gather additional eligibility information, history of psychological or neurological diagnoses, and prior experience with music training. Specifically, parents are asked, *“Has your child participated in any music training program for more than six months?”* Children whose parents answer “yes” to this question are excluded from participation in all study arms. Finally, eligible children complete a remote Zoom assessment with a trained research assistant to assess intellectual functioning using the Wechsler Abbreviated Scale of Intelligence–Second Edition (WASI-II). To ensure that all participants can meaningfully engage with the intervention and that global cognitive delays do not confound observed effects, only children with a WASI standard score above 85 were included. Although primarily a screening measure, it may also serve as a covariate or exploratory variable to examine individual differences in outcomes as a function of cognitive ability.

### Training programs

#### The Music Intervention (Colburn)

The target for participants in the music intervention is to receive a total of 105 minutes of group classes and 30 to 45 minutes of private lessons each week. Each year, we plan that participants also attend a 180-minute retreat per semester and a summer camp (∼10 hours per year). For cohort 1, over the two-year intervention period, the total training duration was approximately 164 hours, including 55 Monday sessions, 61 Friday sessions, 54 private lessons, 22 hours of summer camp, and 12 hours of retreat activities. As cohort 2 is ongoing, final intervention duration data are not yet available.

The lead instructor of the music class holds a BM in music performance and teaching credentials for music education. The instructor has extensive experience teaching youth bands and orchestras at multiple arts organizations across LA, including the Los Angeles Philharmonic’s Youth Orchestra Los Angeles (YOLA) and Colburn School of Music (Colburn). The private lesson instructors are certified Bachelor of Arts students at Colburn Conservatory or School, studying violin performance, or are equivalently certified.

The weekly violin group classes meet twice a week and are taught by the lead instructor with support from the private lesson teachers. All instruction takes place at Colburn School in Downtown LA. The classes focus on building a strong musical foundation in ear training, rhythm, and tension-free violin playing (i.e., posture, motor, and breathing awareness). Classes are structured to incorporate singing, movement, listening, dictation, reading, and performance. Students are frequently given opportunities to build confidence by playing in front of others.

Classes follow the Suzuki Method’s music curriculum and teaching philosophy, Mimi Zweig’s violin pedagogy judgment-free zone, and Paul Rolland’s well-researched methodology of tension-free playing for violinists (Lysaker, 2020; Peak, 2012; Rolland & Colwell, 1966). The repertoire consists primarily of Western classical pieces, children’s songs, scales, arpeggios, and technical exercises for young musicians.

In addition to group class, students receive a weekly 30 to 45-minute private lesson, in which they practice what they’ve learned in the group class. The one-on-one setting of the private lesson allows for closer attention to each student’s technique, intonation, and specific needs, helping them develop their skills and supplement the education in group lessons. Parents, guardians, and siblings are encouraged to observe the group and private lessons so they may encourage and help their child practice at home if needed. The lead instructor creates and shares videos of exercises for students and their families to use and guide at-home practice. These group classes culminate in a final performance for students’ families at the end of each semester.

#### The Theatre Program (24th Street)

The theatre intervention takes place at a renowned, well-established community-based theatre school in Los Angeles. Participants in the theatre intervention attend three group classes a week (Monday, Tuesday, Wednesday) and a summer camp (∼10 hours per year) led by a lead teacher and up to two teaching assistants. The classes are 50 minutes each session, totaling 2 hours and 30 minutes of group instructional time per week. For cohort 1, over the two-year intervention period, the total training duration was approximately 164 hours (174 sessions; summer camp: 19 hours). Three lead teaching artists instruct the students, one on each class day. One assistant is present for every class for continuity between the teachers. The lead instructors hold BFAs in dramatic arts and are active actors, producers, and directors in the LA theatrical community. All instructors have extensive experience teaching youth acting classes and directing youth actors and performers at multiple arts organizations across the city, including Colburn (Drama department), University of Southern California, and the LA Philharmonic. The assistant teachers both have degrees in theatre performance and several years of experience teaching young people in theatre.

Each class begins with a warm-up, moves towards improvisational scenes, and ends with a ritual (which changes depending on the teacher). Scene content includes real-life locations and scenarios, but occasionally ventures into the fantastical. In the improvisational scenes, students and instructors work together to create and enact different imagined scenarios. Additional goals for each class include building confidence onstage and practicing creative thinking on the spot. Topics covered in these beginning classes include tableaux, improvisation, vocal projection, proper diction, pantomime, gesture, character development, and scene setting. Over the first year, the theatre classes focus first on moving on stage, then on adding voice skills, culminating in a final show performed for the students’ families at the end of each semester. In the theatre intervention, background music is occasionally used during performances to support the narrative, but participants are not instructed to synchronize their movements or speech with it. While spontaneous coordination may occur during improvisation, synchronization is not a systematic or intentional component of the training or performance.

### Outcomes

#### Primary Outcomes

The primary objective is to examine the effects of the music training intervention on brain inhibitory control circuitry after two years of participation in the randomized controlled trial (RCT). Inhibitory control will be assessed using behavioral tasks (Go/No-Go and delay gratification) and fMRI. The primary outcome measure is the percent signal change in the ROIs, evaluated separately during the inhibitory control tasks. Specifically, we will analyze the contrast between No-Go and Go in the Go/No-Go task and between delayed-choice and control-choice trials in the delay gratification task. Primary outcomes will be measured at baseline and at the 2-year follow-up.

#### Secondary Outcomes

Secondary outcomes include behavioral indices of inhibitory control. They will be obtained at baseline and at the 2-year follow-up: Go/No-Go signal detection (d’) and the discounting rate from the delay gratification task.

#### Exploratory Outcomes

Exploratory analyses will evaluate the intervention’s effects on additional behavioral and cognitive domains of child development. These outcomes will be collected at baseline, 1-year post-randomization, and 2 years post-randomization (see Table 1 for details) and include:

a. Cognitive ability: NIH Toolbox Flanker task, Wechsler Abbreviated Scale of Intelligence – II (WASI-II).
b. Socio-emotional functioning: Parent BASC, Child Behavior Questionnaire, NIH Toolbox Emotion Battery.
c. Musical skills: MBEMA, BAT, Child Musicality Questionnaire, Goldsmiths Musical Sophistication Index – Child.
d. Motor skills: NIH Toolbox Pegboard.
e. Auditory ability: NIH Words-in-Noise Test.
f. Structural and diffusion neuroimaging measures

**Table 1.**
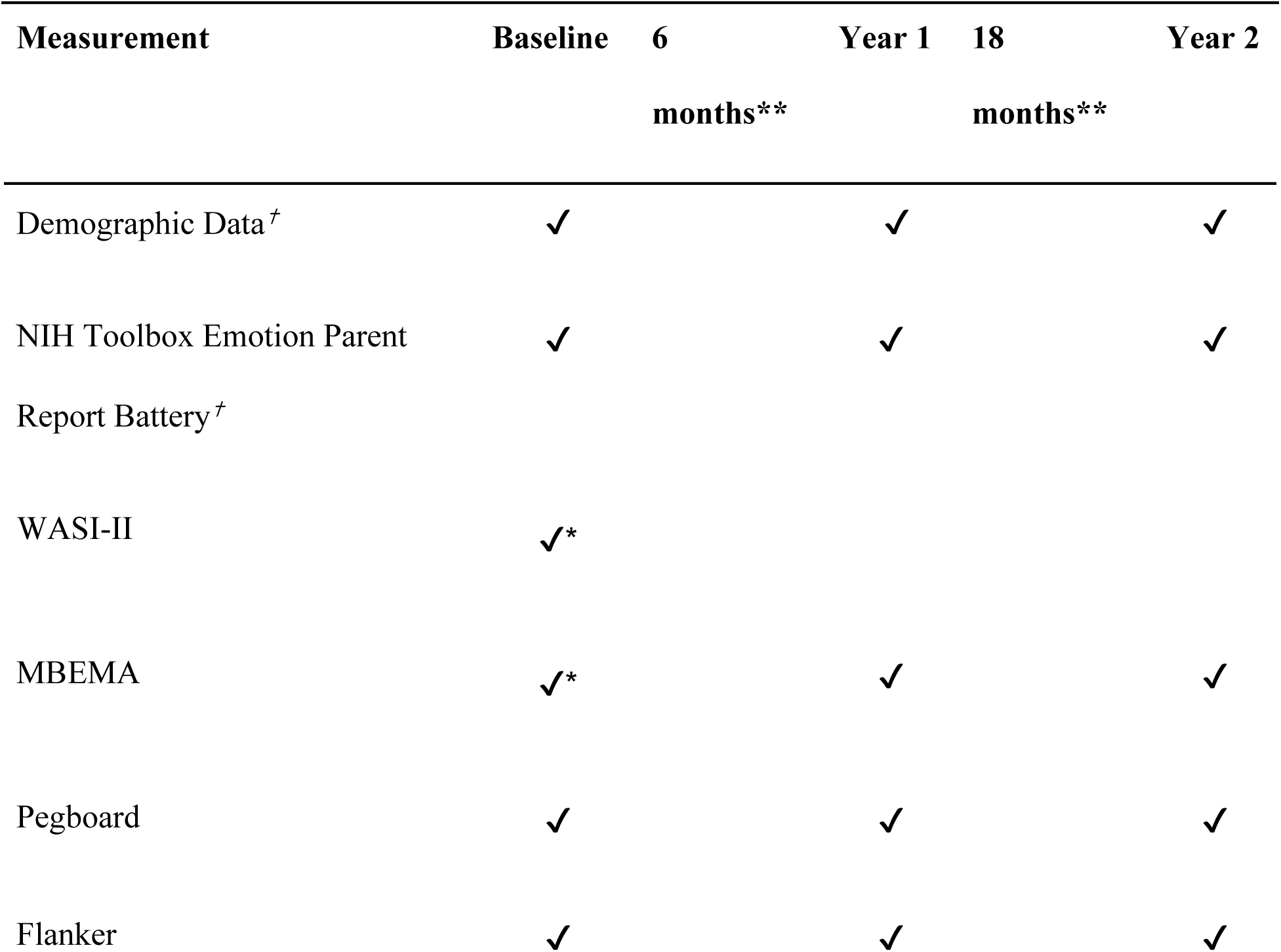
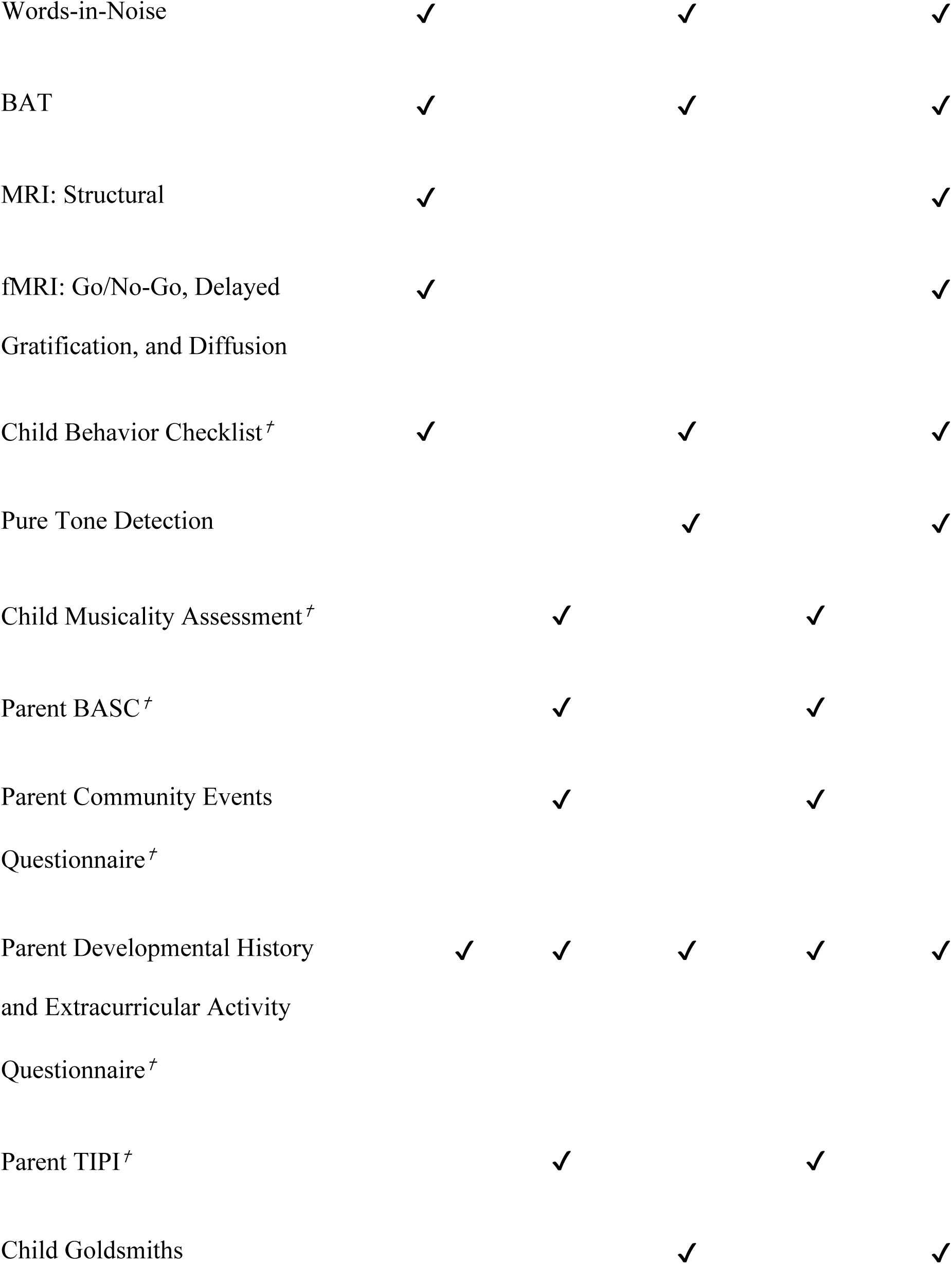
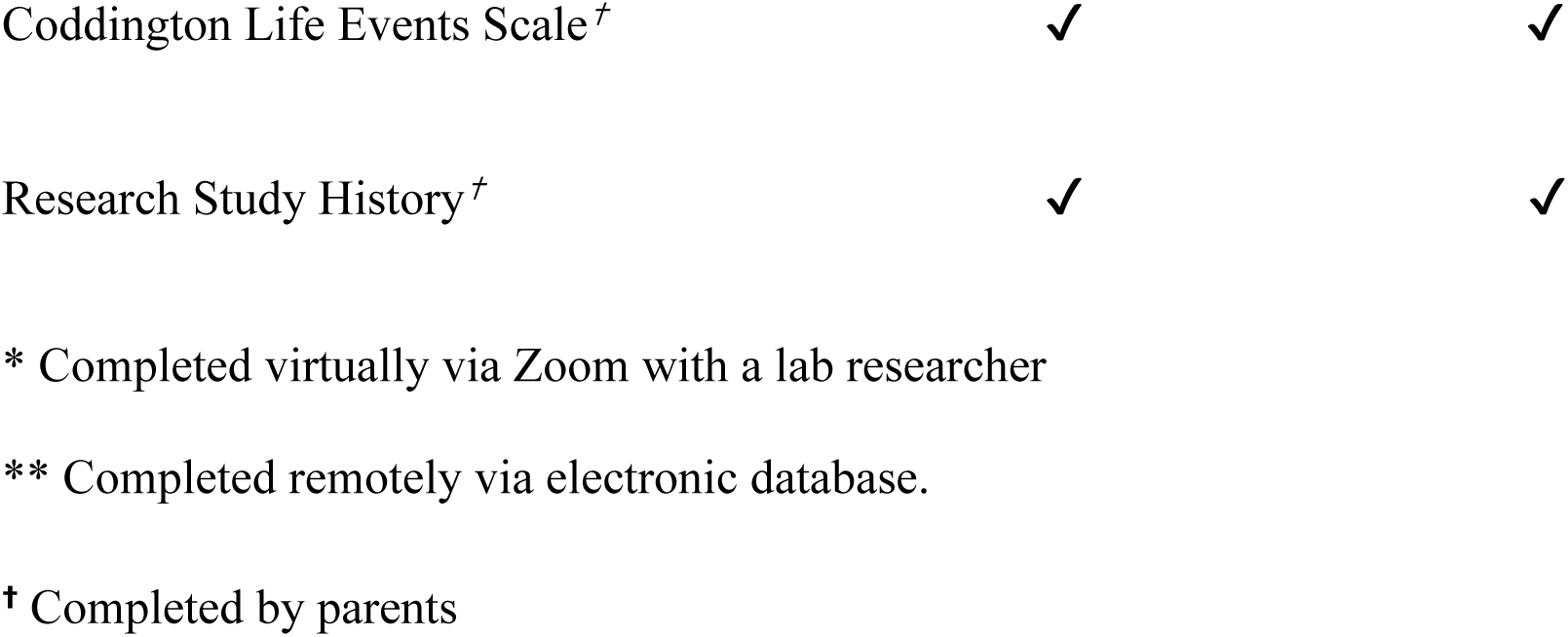
Measurements and Testing Schedule.

### Behavioral Assessments and Measures

An assessment protocol consisting of cognitive, behavioral, and psychological measures, and informative questionnaires (e.g., physical health, home environment, musical activity) designed for the EXCEL trial is used to assess behavioral changes during the protocol. After screening, participants and their parents complete questionnaires and behavioral tests (perceptual, musical, and cognitive) at baseline (pre-intervention), year 1 (mid-intervention), and year 2 (post-intervention). Baseline testing is split into two sessions: one remote (baseline-Zoom) and one in-person (baseline-lab). MRI is completed at baseline and year 2. In addition, mid-trial evaluations using questionnaires are conducted every 6 months via a secure electronic platform. These assessments are used to evaluate changes in the home environment, housing, physical and mental health, engagement in enrichment programs outside the assigned program in the EXCEL trial, and satisfaction with the programming provided. These are followed up by phone calls if necessary. Assessors at the year 1 and year 2 intervention sessions are blind to group allocation. All measures are listed in Table 1, along with the visit schedule, and are described in detail below.

#### Social, Demographic, and Lifestyle Measures

##### Coddington Life Events Scale–Child (CLES-C)

The CLES-C for children ages 6 to 12 years is a 36-item questionnaire that collects information on major life events (LE) and their potential impact on the child’s life, growth, and adjustment (Coddington et al., 1999).

Respondents indicate for each LE the number of times the event occurred in the last 12 months. Responses are weighted based on the amount of time since the event occurred and the stress associated with the given event into Life Change Units (LCUs); a total LCU score is obtained for each participant to indicate the relative impact these events had on the individual’s growth and adjustment over the last 12 months.

##### Parent Developmental History and Extracurricular Activity Questionnaire

A modified version of the Family History Interview developed by Ilari et al. (2019) is administered to assess (1) demographics and family life, (2) medical history of the child and family, (3) language information, (4) schooling, (5) engagement in extracurricular activities, and (6) the child’s interest in music and theatre activities. Items 1-4 are collected once at baseline, and items 5 and 6 at all subsequent time points.

##### Parent Community Events Questionnaire

Modeled on the scale used in Ilari et al. (2019), this six-item questionnaire assesses the frequency of witnessing various forms of community violence. Parents answer as a proxy for their child to indicate how often their child experienced various events (options are never, once, twice, or more than twice).

##### NIH Toolbox Emotion Parent Report Battery

To assess children’s emotions, parents complete the Emotion Parent Report Battery (Gershon et al., 2013). Three subcategories are *affect* (anger and sadness scales), *well-being* (positive affect and general life satisfaction scales), and *social relationships* (social withdrawal, positive peer interaction, peer rejection, and empathetic behavior scales). Each scale consists of 5-12 statements. Parents rank each item using a 5-point Likert scale.

##### Research Study History

A short questionnaire detailing families’ previous experience with research and their comfort level with fMRI is better for understanding our population’s experience with scientific research.

#### Cognitive, Behavioral, and Psychological Measures

##### Wechsler Abbreviated Scale of Intelligence, Second Edition Full Scale Intelligence Quotient-2 Subtest (WASI-II; FSIQ-2)

To measure overall cognitive ability, the WASI-II (2-subset test) is administered at baseline (Wechsler, 2011). This standardized intelligence assessment consists of 2 subtests: Vocabulary and Matrix Reasoning. The Vocabulary subtest consists of up to 31 questions designed to assess an individual’s verbal abilities and vocabulary skills. The assessor orally presents words and asks the child to define each. The Matrix Reasoning subtest consists of up to 30 questions designed to assess an individual’s fluid intelligence, visual intelligence, spatial abilities, classification skills, and perceptual organization skills. Children are presented with sets of images and asked to choose one to complete a pattern or a set. The two subtests provide a combined intelligence quotient (FSIQ-2).

##### NIH Toolbox Nine-Hole Pegboard Dexterity Task – V2

The measurement of fine motor manual dexterity is evaluated with the 9 Hole Pegboard Dexterity Task (Gershon et al., 2013). Participants are assessed on their speed for placing and removing nine pegs into a pegboard, once with each hand (dominant and non-dominant).

##### NIH Toolbox Flanker Inhibitory Control and Attention Test (Flanker) – V2

The Flanker test measures a participant’s attention and ability to inhibit automatic responses that could interfere with achieving goals (Gershon et al., 2013). Participants are presented with five arrows in a row and asked which direction the *middle* arrow points (left or right). On congruent trials, all arrows are facing the same direction as the middle arrow, whereas on incongruent trials, the flanking arrows are facing the opposite direction from the middle arrow. The performance difference between congruent and incongruent trials reflects the cognitive cost of resolving conflicting information. A single composite score is calculated that accounts for trade-offs between speed and accuracy (LaForte et al., 2024). Participants under the age of 8 are administered a practice task using fish in place of arrows. During the EXCEL trial, NIH Toolbox V3 was released, which slightly altered some assessments, such as removing the fish from the practice task for younger participants. The EXCEL trial continued to use NIH Toolbox V2 for consistency throughout data collection.

##### Child Behavior Checklist (CBCL)

A measure of emotional and behavioral difficulties in children. This parent-completed 113-item checklist scores items on a 3-point Likert scale and consists of 8 syndrome scales (anxious/depressed, depressed, somatic complaints, social problems, thought problems, attention problems, rule-breaking behavior, aggressive behavior) and two higher-order factors (internalizing and externalizing; Achenbach, 1991). In addition to these questions, parents also complete competence scales for socializing, activities, school, and total competence.

##### The Behavior Assessment System for Children Parent Rating Scale (BASC-2 PRS)

This 30-item questionnaire assesses a child’s behavior and emotional adjustment from the parent’s perspective (Reynolds, 2010). Parents answer each question on a 4-point Likert scale: “Never”, “Sometimes”, “Often”, or “Always”. This scale includes questions to assess the child’s activities of daily living, adaptability, aggression, anxiety, attention problems, conduct problems, depression, functional communication, hyperactivity, leadership, social skills, and somatization.

##### Ten Item Personality Inventory (TIPI)

This inventory is a 10-item questionnaire designed to assess the Big Five personality traits: extraversion, agreeableness, emotional stability, conscientiousness, and openness (Gosling et al., 2003). Parents complete this scale for their children.

#### Auditory Function Measures

##### NIH Toolbox Words-in-Noise Test – V2

The Words in Noise task measures a participant’s ability to hear words in a noisy environment (Gershon et al., 2013). Participants are asked to pick out the sound of a target female voice asking them to say certain words among different levels of background babble talker noise. As the participant hears each word from the target voice, they are instructed to repeat it aloud. The task consists of 7 rounds with different signal-to-noise ratios: 24, 20, 16, 12, 8, 4, and 0 dB. The task is discontinued if the participant fails to repeat all five words in each round. This task is repeated for both the right and left ear.

##### Pure Tone Detection

The pure tone detection threshold task is based on a standardized AMCLASS audiogram to collect audiometric thresholds bilaterally at 500, 1000, 4000, and 8000 Hz and is run on the TeamHearing.org platform (Goldsworthy, 2020). The AMCLASS audiogram classification uses standardized rules to categorize configuration, severity, and intramural asymmetry of an audiogram, allowing descriptions of different types of hearing loss (Margolis & Saly, 2007). Participants are asked to discern the pairing of a visual stimulus with a tone. Trials decrease in volume until a lower threshold is measured for 400 ms sinusoids with 20 ms raised-cosine attack and release ramps at each of the 7-octave frequencies from 110 to 7040 Hz. Participants are given three alternatives in a three-interval forced choice method: three buttons labeled “1”, “2”, and “3” appear on the screen. For each trial, the buttons flash in numerical order, and one of the three flashes is paired with a tone starting at the participant’s reported “medium” loudness level (assessed before the task). In contrast, the other two buttons are paired with silence. The button tone is matched and varied across trials. Following the button flashes, the participant chooses the button paired with the tone. Correct responses result in 1-step gain reductions, while incorrect responses result in gains increased by three times the step in the subsequent trial. The first step is 6 dB and is reduced by 2 dB after the first correct answer following an error. Participants are allowed four errors in the task; after that, the run concludes, and the final gain value is taken as the lower threshold for tone detection. This procedure is repeated for each of the four frequencies until all thresholds are obtained.

#### Measures of Musical Skill

##### Montreal Battery of Evaluation of Musical Abilities (MBEMA)

Tonal and rhythmic aptitudes and musical memory abilities are measured using the MBEMA (Peretz et al., 2013). In the first two sections, the participant is required to compare two unfamiliar melodies and determine whether they are the same or different. In the tonal section, the melodies may differ in their melodic contour. In the rhythm section, the melodies may vary in their rhythms. In the melodic memory section, participants are asked whether they can recognize melodies from the first two sections by distinguishing them from brand-new melodies.

##### Beat Alignment Test (BAT)

The BAT measures beat-based processing abilities (Iversen & Patel, 2008). Part 1 assesses sensorimotor synchronization—the ability to align motor movement (finger tapping) with an internal (preferred tempo) or external stimulus (e.g., a musical beat or metronome). Participants are asked to tap a steady beat at their preferred tempo and then complete paced-tapping tasks with a metronome for 30 seconds each at IOIs of 400ms, 550ms, and 700ms. In part 2, participants are asked to tap along to the perceived beat of 12 different music clips. Accuracy and the regularity of tapping are assessed using circular statistics (vector direction) and inter-tap intervals. Participants are also asked about their familiarity with the musical stimuli. Lastly, part 3 assesses beat perception: participants listen to music clips with a series of beeps and are asked whether the beeps align with the music’s beat.

##### Child Musicality Assessment Inventory for Teachers and Parents

This 9-item questionnaire assesses a child’s musical skill set through teacher or parent report (D. Müllensiefen & E. Hannon, personal communication, 2022a) and comprises three factors: Perception, Production, and Enthusiasm for Music.

##### Goldsmiths Music Sophistication Index–Child (Gold-MSI-C)

The *Gold-MSI-C* is a child-adapted self-report version of the original Goldsmiths Musical Sophistication Index (D. Müllensiefen & E. Hannon, personal communication, 2022b). It includes 13 items scored on a 5-point Likert scale, assesses enthusiasm for music and music making, and calculates a global score. Additional data on musical training and pitch perception (a single item) are also collected, but are not part of the factor structure.

### Neuroimaging Measures

Children complete structural (T1-weighted MPRAGE), diffusion, and functional MRI scans. A T2-weighted sequence is also acquired and reviewed by a neuroradiologist for clinical purposes. In the event of an incidental finding, the neuroradiologist contacts the family-designated physician and recommends follow-up if warranted. All MRI data are acquired using a 3T MAGNETOM Prisma scanner with a 32-channel head coil.

An MRI protocol adapted for children is implemented, including a training session before scanning. They are first familiarized with the scanning environment using a mock scanner, exposure to scanner sounds, and practice in remaining still. During the MRI session, the child may choose to be accompanied by someone in the scanning room. If selected, an investigator remains in the scanner room to provide reassurance and reminders. Participants may bring an MRI-safe stuffed animal for comfort. At the start of each in-person visit, children select a “Squishmallow” from the lab’s collection to serve as their “buddy” for the day, which can also accompany them into the MRI scanner. To minimize motion during structural and diffusion acquisitions, children watch a video of their choice from a streaming platform. They are covered with a weighted blanket throughout the session, including functional sequences.

Real-time motion estimates during functional scans are obtained using FIRMM (Framewise Integrated Real-Time MRI Monitoring) software to ensure data quality. Scans are considered acceptable if more than 70% of volumes exhibit framewise displacement (FD) < 0.4 mm. If this threshold is not met, the scan is stopped, and participants are reminded to remain still. When possible, the sequence is restarted to improve data quality. Upon completion of the session, participants receive a printed image of their brain.

#### Structural imaging

High-resolution T1-weighted structural MR images are acquired using an MPRAGE sequence with the following parameters: 1 mm × 1 mm × 1 mm resolution over a 256 mm × 256 mm × 256 mm FOV; TI/TE/TR = 1060/2.82/2500 ms; flip angle = 8 degrees;° GRAPPA acceleration factor *R* = 2.

#### Functional imaging

Functional images are obtained using a gradient echo, echo-planar, T2-weighted pulse sequence (TR = 800 ms, one shot per repetition, TE = 30 ms, flip angle = 52,° 90 x 90 in-plane resolution). Sixty slices covering the entire brain are acquired with a voxel resolution of 2.4 mm x 2.4 mm x 2.4 mm over a 216 mm x 216 mm x 216 mm FOV. Three hundred and thirty-eight volumes are collected during each run of the Go/No-Go task, and up to 750 volumes are collected during each run of the delayed gratification task. Task order is randomized. To process fMRI data, FSL (Smith et al., 2004) is used with standard preprocessing (i.e., fMRIprep; Esteban et al., 2019) and GLM modeling. Group-level comparisons use cluster correction (z > 3.1, p < 0.05). T1 images are processed using BrainSuite and FreeSurfer v6.0 for cortical thickness and volumetric segmentation. Thickness is computed using PVC and averaged across ROIs. Manual quality check is performed on all segmentations.

#### Go/No-Go

This fMRI task is a child-adapted Go/No-Go paradigm modeled after Réveillon et al. (2013) to assess inhibitory control (Figure 2A). Each trial begins with the presentation of a colored spaceship (green or red). Participants are instructed to press a button as quickly as possible for green spaceships (“go” trials) and to withhold responses for red spaceships (“no-go” trials). Each of the two functional runs includes 30 go trials and 10 no-go trials, for a total of 40 trials per run. Trials begin with a jittered fixation cross (M = 2900 ms, SD = 928 ms), followed by the stimulus (500 ms) and an interstimulus interval (2500 ms). Whether or not a response is made during a trial, the interstimulus interval is 2500–3000 ms to maintain consistent trial durations. Each run lasts approximately 240 seconds (300 TRs). Accuracy (percent correct) is recorded for each trial type, and reaction times are collected for go trials and commission errors on no-go trials.

**Figure 2A.**
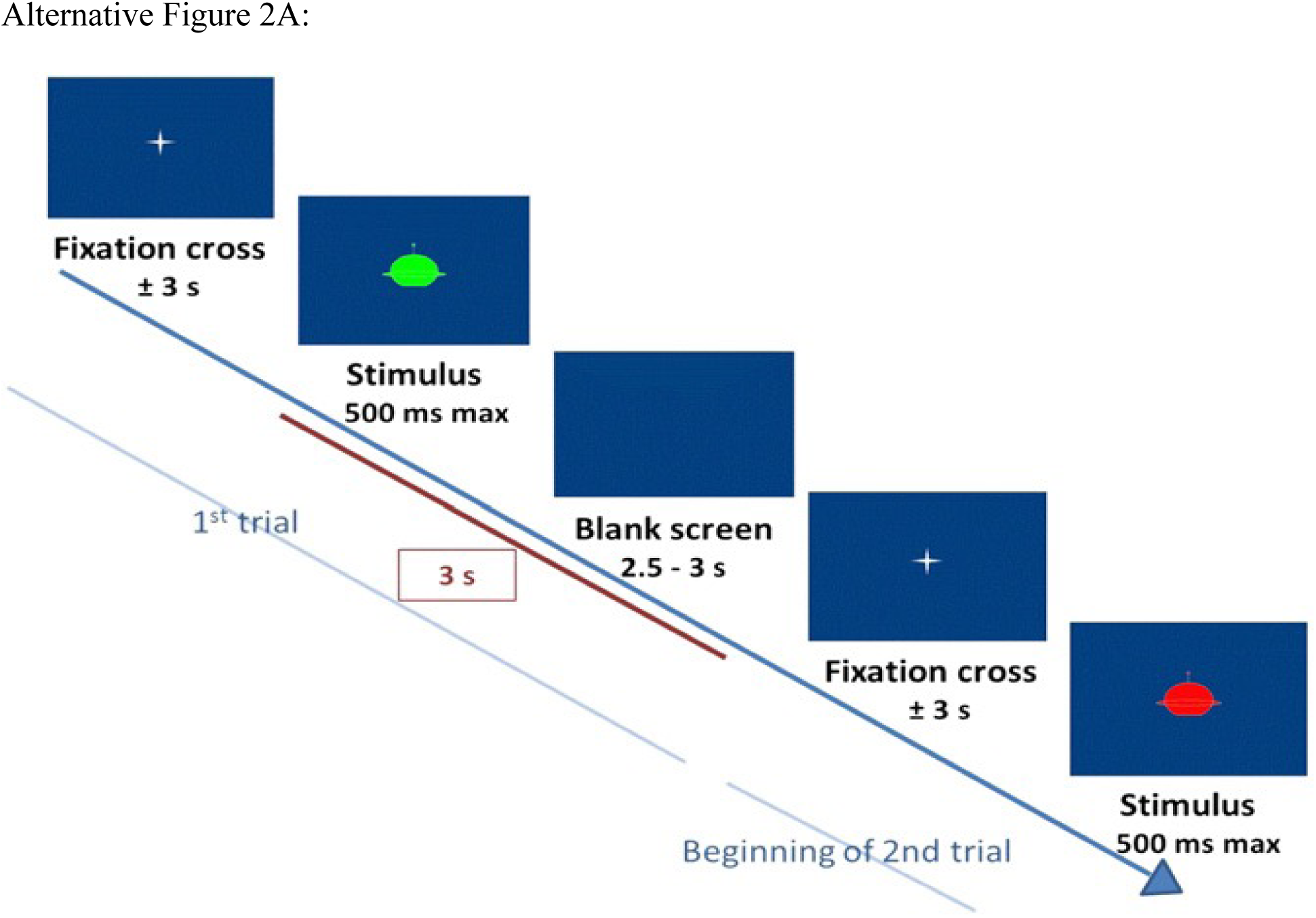
Go/No-Go Task

#### Delayed Gratification

This fMRI task is a modified version of a paradigm developed by Scheres et al. (2014) to assess delayed gratification in children aged 6–12 years (Figure 2B). In each trial, participants choose between two airplanes carrying varying amounts of coins (2, 4, 6, 8, or 10 cents) that fly from opposite sides of the screen. Airplanes positioned higher in the visual field indicate a longer delay before reward delivery compared to those positioned lower. Trials are divided into three conditions. In the main condition (1), participants choose between a smaller reward delivered sooner and a larger reward delivered later. In the two control conditions, either the coin amounts differ while the delay remains constant (condition 2), or the delay differs while the coin amounts remain equal (condition 3). These control trials serve as a baseline for fMRI analysis and verify task comprehension. Selection of the inferior option in control conditions (i.e., choosing less money or longer delay with no added value) on more than 30% of trials results in exclusion from analysis, as this suggests inattention or misunderstanding. Participants complete two functional runs of 20 trials each (16 from condition 1, 2 each from conditions 2 and 3). During practice, children are informed that they can exchange their accumulated coins for toy prizes.

**Figure 2B.**
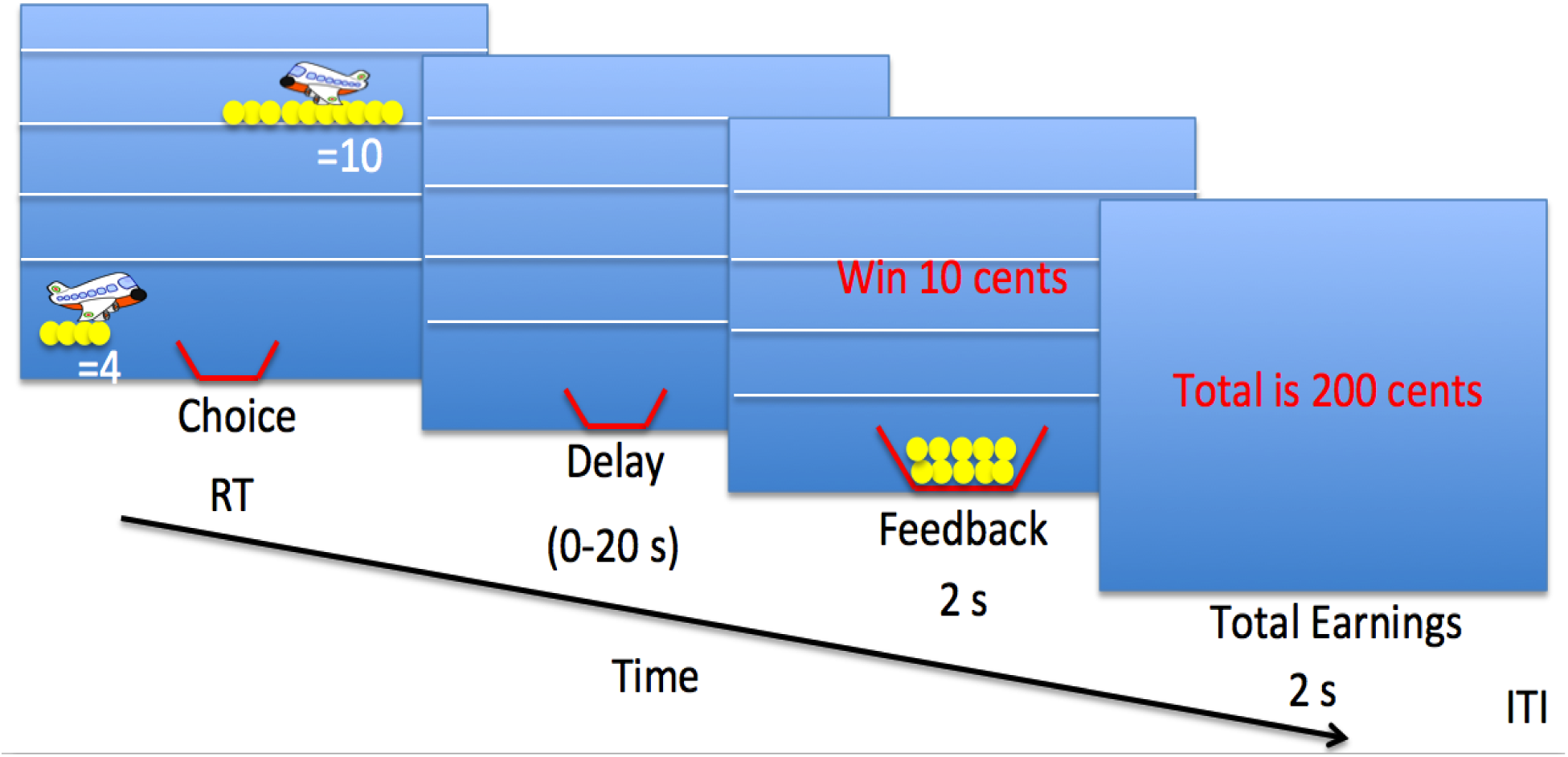
Delayed Gratification Task

Each trial begins with stimulus onset, during which participants have up to 10 seconds to select their preferred option by pressing the corresponding button (left or right). Stimuli remain on screen until a response is made or the 10-second window elapses, after which the trial proceeds automatically. Based on the chosen stimulus and trial condition, a variable “delay” period (0, 5, 10, 15, or 20 seconds) follows, during which the screen remains static. After the delay, coins are visually delivered from the airplane to the participant’s virtual basket at the bottom of the screen. This is followed by two feedback displays: the number of coins gained (2 seconds) and the updated cumulative total (2 seconds). Delay durations are not explicitly shown; instead, participants experience each delay interval during practice, allowing them to associate airplane height with time-to-reward. An interstimulus interval of 0.5 to 2 seconds separates consecutive trials to optimize event timing for fMRI analysis. Run durations vary depending on participant response speed and reward selection, typically ranging from 180 to 600 seconds (225–750 TRs). Reaction time and accuracy are recorded. Responses are coded as delayed or immediate in condition 1, and as superior or inferior in conditions 2 and 3.

#### Diffusion

Diffusion MRI data is acquired using one image without diffusion weighting (b= 0 s/mm^2^) and 30 diffusion-weighted images (b= 1,000 s/mm^2^) with orientations distributed uniformly over the sphere. Acquisitions use a standard twice-refocused spin-echo EPI pulse sequence with the following parameters: 2 mm x 2 mm in-plane resolution over a 260 mm x 260 mm field of view, 66 slices with 2 mm slice thickness and no slice gaps, TE/TR = 80/9,000 ms, and 6/8ths partial Fourier encoding with no parallel imaging acceleration. Rather than using the images produced by the scanner, images are directly reconstructed using custom in-house methods. To eliminate Nyquist ghosts, we estimated linear-phase correction factors to account for mismatches between k-space lines acquired with opposite gradient polarities.

Phase correction factors were obtained using variable projection methods (Haldar et al., 2007) to optimize a cost function with two components:

1. A least-squares penalty on the mismatch between phase-corrected even and odd navigator lines acquired without phase encoding gradients (Bernstein et al., 2004).
2. A nonconvex penalty, given by the Schatten-*p* seminorm (*p* = 0.5), applied to a Toeplitz-structured matrix formed from the combined phase-corrected even and odd k-space lines (Lobos et al., 2018).

To reduce noise, images are then reconstructed jointly from the corrected k-space data using SNR-enhancing joint reconstruction (Haldar et al., 2013), with sensitivity maps estimated using ESPIRiT (Uecker et al., 2014) with PISCO accelerations (Lobos et al., 2018; Lobos & Haldar, 2022) and phase constraints estimated from low-frequency k-space data. Following reconstruction, a B0 field map is calculated from the reversed gradient data using FSL’s top-up software (Andersson et al., 2003), and geometric distortion is addressed using spherically regularized distortion correction (Bhushan et al., 2014).

### Statistical Methods

Planned analyses include feasibility outcomes and comparison of randomized interventions on changes in IC for behavioral and neuroimaging outcomes.

#### Sample Size Estimation

The primary outcome of the EXCEL trial is activation of prefrontal inhibitory control networks during fMRI tasks. Based on prior studies of the effects of music training on children (Hennessy et al., 2019; Zuk et al., 2014), effect sizes in key ROIs ranged from 0.69 to 1.47, with a median of 0.85. Using a conservative estimate (d = 0.8) for sample size calculation, and accounting for clustering within 16 intervention groups (ICC = 0.025), 85% power, and 20% attrition, the EXCEL trial target was to randomize 114 participants (57 per condition; 7–8 per group). A total of 129 participants were randomized. For details on the flow of participants, see Figure 3.

**Figure 3.**
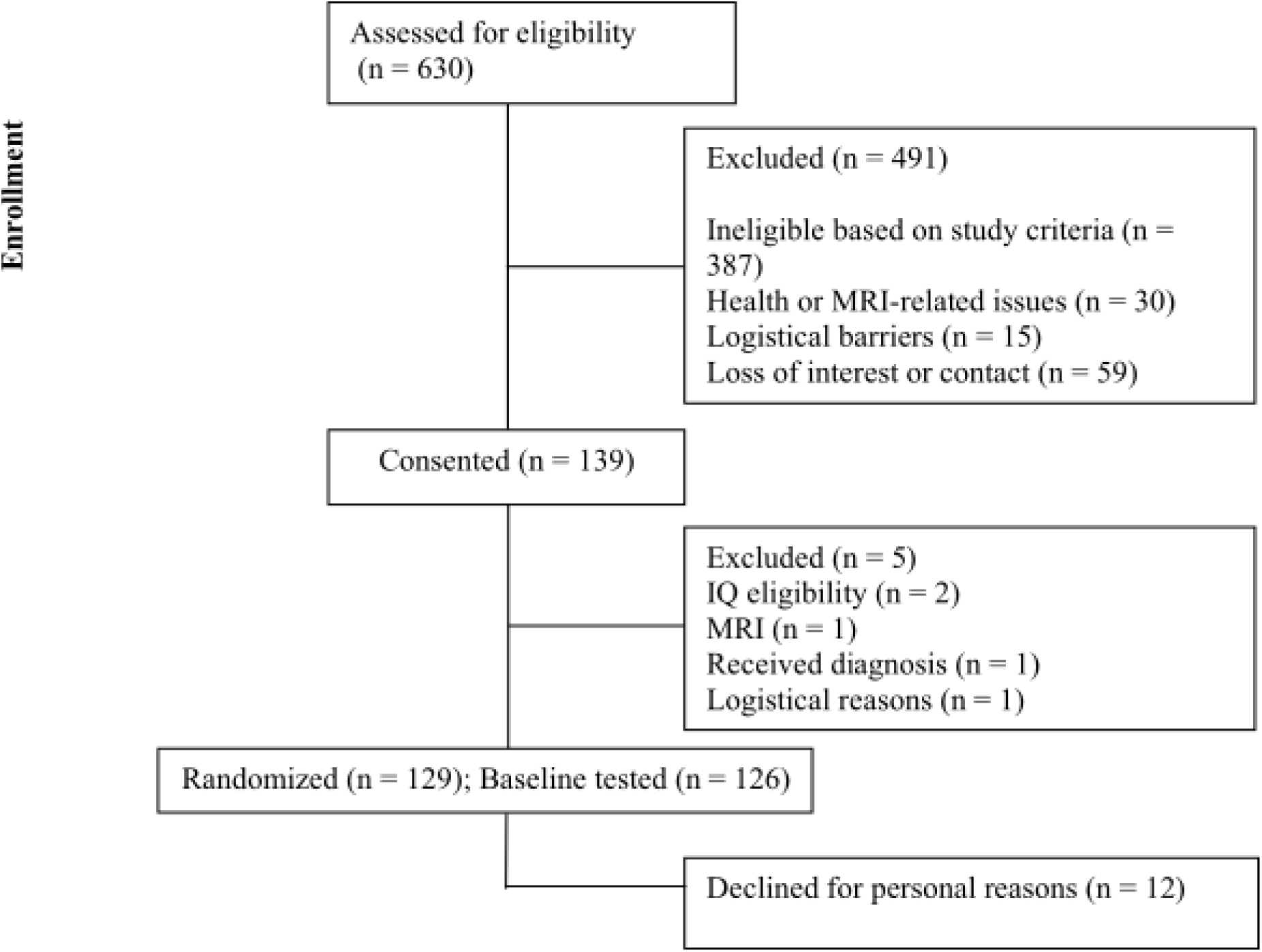
CONSORT Flow diagram of participants through phases of the RCT.

#### Outcomes

##### Feasibility

Assessed through retention (≥70%), adherence (≥70% session attendance per participant), and continuation intent after one year.

##### Primary Outcomes

Go/No-Go performance is assessed via accuracy on go/no-go trials. For fMRI, the primary contrast for Go/No-Go is correct no-go vs. go trials; for delayed gratification, delayed vs. control choices. Activation in the ROIs is extracted based on a separate dataset. Functional connectivity analyses (PPI) examine task-modulated coupling between these regions and limbic areas (striatum, amygdala). Primary analysis will be by intention-to-treat, in which participants are analyzed according to their randomized intervention regardless of adherence. Linear mixed-effects models will be used to assess intervention effects on trial outcomes, with random intercepts for participant and ROIs. Main effects of group, time (year 2 vs. baseline), and the group × time interaction will be tested for fMRI signal changes in ROIs during each IC task. Covariates will include randomization stratification factors (age, sex) and any baseline imbalances (standardized difference ≥0.1). A per-protocol sensitivity analysis will exclude non-adherent participants (> 30% adherence). A second sensitivity analysis will use multiple imputation (by chained equations) for missing outcome data.

##### Secondary Outcomes

Delayed gratification is indexed by discounting rate, calculated as area under the curve (AUC) using the trapezoidal method. Secondary behavioral outcomes, including d’ (Go/No-Go) and the discounting rate (delay task), will use the same statistical approach (without ROI nesting).

##### Exploratory outcomes

We will apply linear mixed-effect models to test for intervention-group differences in changes in cortical thickness, subcortical volume, and prefrontal-limbic connectivity across the whole brain and ROIs identified in previous research (Habibi, Damasio, Ilari, Veiga, et al., 2018). Mixed models assess additional behavioral measures, including effects on emotion (BASC, NIH Emotion Toolbox), cognition (WASI-II; NIH Toolbox Flanker), musical ability (MBEMA, BAT, Goldsmiths-Child), motor function (NIH Pegboard), and auditory function (Words-in-Noise, pure-tone detection).

### Interim Analyses

No formal interim analyses for efficacy are planned. However, feasibility outcomes (e.g., retention, adherence, and fidelity) are evaluated at predefined timepoints (e.g., 12 months and study completion) to assess study progress and inform continuation to subsequent phases of the trial.

### Establishing Partnerships: Intervention Delivery

In June 2022, we initiated outreach to 35 reputable after-school programs in the greater downtown Los Angeles area via email and phone to assess interest in partnering on a research study examining the impact of music and arts programming on 6- to 8-year-old children. Fifteen programs responded and participated in preliminary conversations focused on their experience serving communities with children from diverse backgrounds, the available infrastructure for two years of weekly programming, and their willingness to foster a welcoming environment for families new to arts education. Following this process, four programs were selected for final interviews based on their location, long-standing presence in the community (10+ years), and demonstrated commitment to underserved communities.

For the music intervention, we partnered with the Colburn School, an internationally recognized performing arts institution in the heart of Downtown Los Angeles. The Colburn Conservatory is acclaimed for cultivating the next generation of professional musicians. At the same time, the Community School of Performing Arts was chosen for its strong commitment to accessibility and excellence, offering high-quality music education to students from across the broader Los Angeles community.

Using an active arts-based control intervention (theatre) rather than a no-activity group allows us to differentiate the specific effects of the music intervention from the general benefits of structured artistic engagement. For the active control group, we partnered with *24th Street Theatre Company*, a nonprofit, award-winning community arts organization that has served the local population for over 25 years. 24th Street Theatre offers theatre-based after-school programming that is specifically designed to reflect and engage the surrounding community.

To ensure smooth coordination and communication with families and program staff, one study coordinator is designated as aware of group assignments (“unblinded”). This individual maintains ongoing contact with participating families and program sites, facilitates logistics, and responds to needs throughout the EXCEL trial. A second team member, unaware of the intervention assignment, assists during testing visits to minimize potential bias. To foster family engagement and ensure clear communication throughout the EXCEL trial, we establish close collaboration between research staff and both partner organizations through regular coordination meetings, emails, and phone calls. Equally consistent communication via text messaging and phone takes place between the unblinded coordinator and participating families. To foster accountability and sustain motivation, each semester concludes with a showcase in which participants present their work, either a musical or theatre performance, depending on their assigned group. Additionally, a group messaging application is used to help families connect and build community, and is overseen by the unblinded coordinator.

### Program Administration and Participation: Fidelity and Adherence

The fidelity of the after-school programs is tracked through two weekly surveys, administered via a secure web-based portal by external observers and the instructional team, who send the research team lesson plans and engagement data. The fidelity data collected by both the teachers and the observers includes (1) the day’s lesson plan and teaching practices, (2) content consistency with planned lesson activities, (3) unanticipated events and student behavior, and (4) if musical activities such as singing or synchronized dancing took place (in the theatre group only). In addition, the observer fidelity measure assesses (1) time spent working independently, in small groups, or in large groups, (2) class engagement level, (3) teacher engagement level, and (4) time spent on essential content areas during each class period.

For music, observers assess the amount of time spent on (1) pitch training (including intonation, ear training, pitch discrimination and singing), (2) rhythm training (including rhythm guided motor control/body movement, clapping or stomping to a beat, playing rhythm with precision instruments, or rhythmic synchronizing with others), (3) sight reading, and (4) social engagement/interaction/attention to rules (including following teacher instructions, taking turns playing speaking and engaging, and socializing with other students). For theatre, observers assess the amount of time spent on (1) perspective taking/role playing (including imagining being someone else and empathy for other people), (2) rhythm guided motor movement (including dancing, moving together in time, clapping/stomping), (3) other music based activities (including singing or playing music), and (4) social engagement/interaction/attention to rules (including following teacher instructions, taking turns playing speaking and engaging, and socializing with other students).

To assess adherence, program teachers record daily attendance at each after-school program and convey it to the research staff through a shared spreadsheet. Research staff code absences according to the reason for the absence (e.g., illness, medical appointment, vacation, family issues), with emergencies or illnesses coded as an excused absence that does not penalize a student’s attendance record. Each month, the research team creates an attendance report for both individual students and the program as a whole to assess adherence to the after-school program. Participants who are present for at least 70% of the month’s classes (our study feasibility benchmark and the attendance requirement for remaining enrolled in the program) are entered into a monthly attendance raffle, where they can win prizes such as movie tickets or a restaurant gift card. The study team contacts participants with attendance below the 70% benchmark for the month to check on well-being and to offer any assistance that can be provided to encourage attendance. They are also reminded of the importance of attendance to remain enrolled in the program and the study. If a child is having trouble getting to the program, public transportation cards are provided to the family to assist with the child’s attendance. Additionally, the research team can help families connect to arrange carpooling.

### Ethics Statement

The EXCEL trial was reviewed by the USC Institutional Review Board (IRB) and was approved by the ethics committee (Approval number UP-22-00056). Parents/legal guardians signed written consent forms, and verbal and written assent were obtained from each child upon enrollment in the EXCEL trial. Children were free to discontinue if no longer interested. Adults received $25 per hour for their child’s participation in research assessments, and the children received tokens they could trade in for prizes (such as small toys or stickers), which they chose upon completing each testing session. At the second and third visits (year 1 & year 2), families received an additional $50. Any important protocol modifications (e.g., changes to eligibility criteria, outcomes, or analyses) will be reviewed and approved by the Institutional Review Board (IRB) at the University of Southern California prior to implementation. Amendments will be documented and updated in the ClinicalTrials.gov registry (NCT05502939) as required. Relevant stakeholders, including study personnel, collaborating institutions, and participants (when applicable), will be informed of significant changes.

### Confidentiality

All participant data will be handled in accordance with institutional and regulatory data protection standards. Identifiable information will be collected only as necessary and will be stored separately from study data using coded study IDs. De-identified data will be used for analysis.

Electronic data will be stored on secure, encrypted, password-protected institutional servers, with access restricted to authorized study personnel. Any physical records will be stored in locked cabinets within secure research facilities. Only approved study personnel will have access to identifiable data. Data sharing, if applicable, will involve only de-identified datasets in compliance with ethical and regulatory guidelines.

### EXCEL trial Progress

Below, we present the demographic characteristics of both participant cohorts (see Table 2), along with a report on retention, adherence, and fidelity for cohort 1, and the progress to date for cohort 2.

**Table 2.**
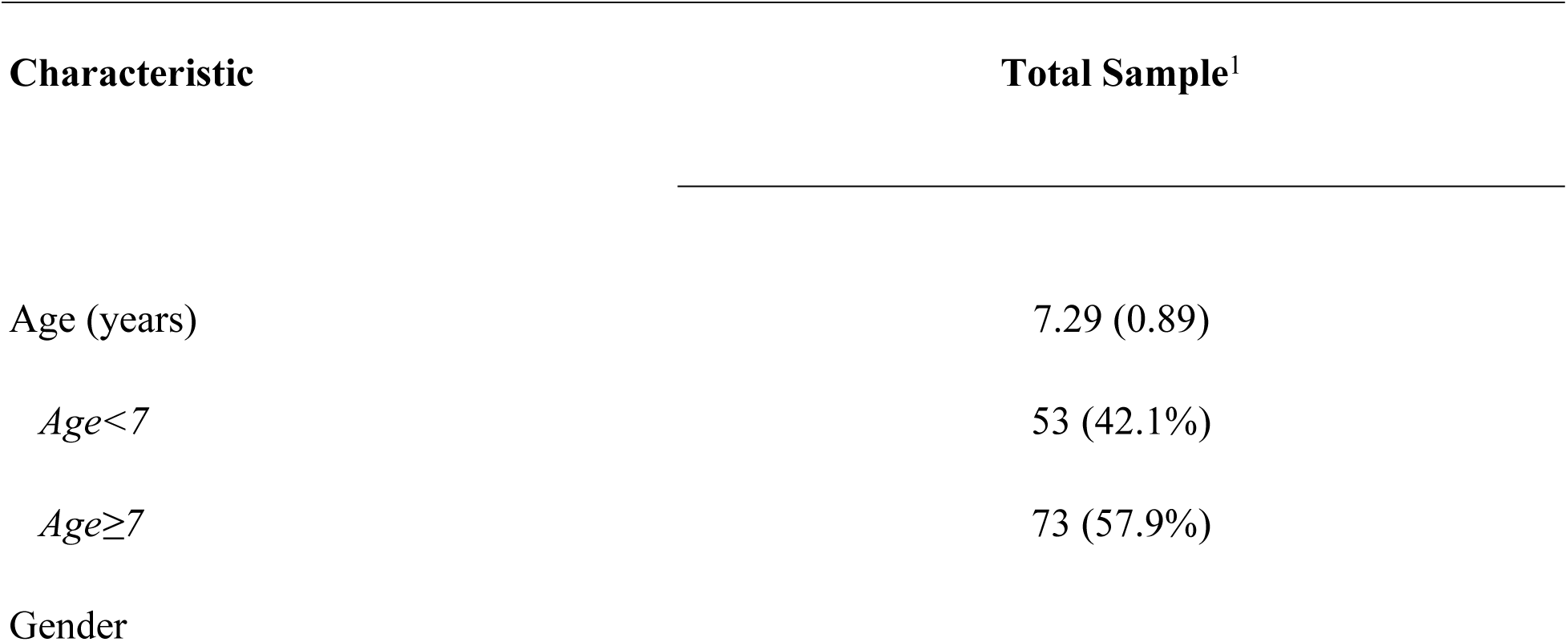
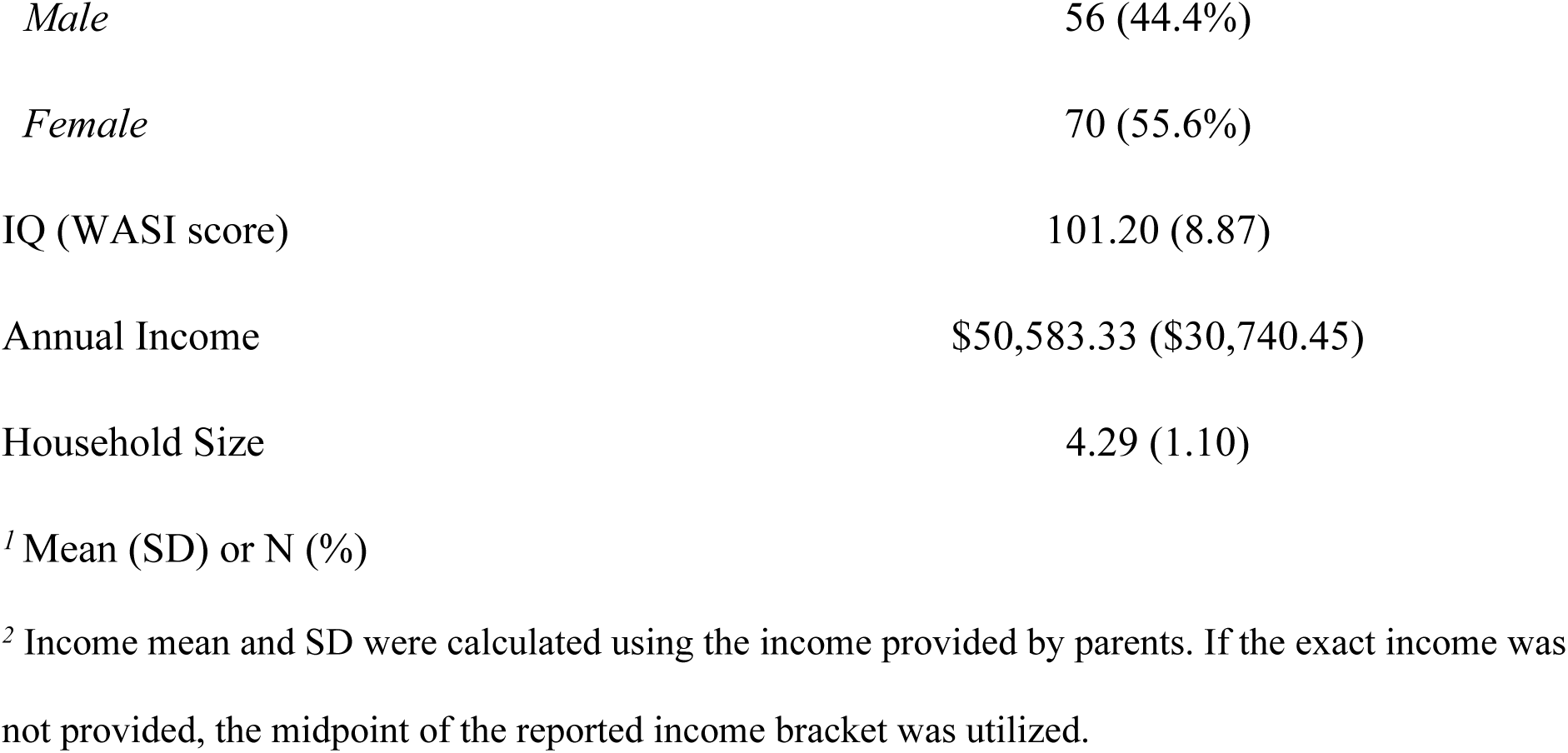
Demographic characteristics of randomized participants (n=126)

| Characteristic | Total Sample<sup>1</sup> |
| --- | --- |
| Age (years) | 7.29 (0.89) |
| Age<7 | 53 (42.1%) |
| Age≥7 | 73 (57.9%) |
| Gender |  |
| *Male* | 56 (44.4%) |
| *Female* | 70 (55.6%) |
| IQ (WASI score) | 101.20 (8.87) |
| Annual Income | \$50,583.33 (\$30,740.45) |
| Household Size | 4.29 (1.10) |

<sup>1</sup> Mean (SD) or N (%)
<sup>2</sup> Income mean and SD were calculated using the income provided by parents. If the exact income was not provided, the midpoint of the reported income bracket was utilized.

#### Cohort 1: Retention, adherence, and fidelity

Retention is defined as the number of randomized and enrolled participants who remain in the program at given timepoints (e.g., year 1 or study completion). Of the 42 students enrolled and randomized, 36 (85.71%) were retained at the one-year mark, and 34 (80.95%) at the end of the EXCEL trial, exceeding the pre-specified feasibility threshold of 70.00%. Adherence is defined as attendance at ≥70% of offered sessions during the first year. Among participants who completed year 1, 34 out of 42 students (80.95%) met the adherence criterion. Average session attendance among retained participants was 92.19%. These results indicate that the EXCEL trial met both pre-defined feasibility criteria for retention and adherence after one year of programming.

In cohort 1, a total of 173 reports were submitted to assess fidelity. Each intervention had different external observers rotating to submit reports (4 observers in year 1 and 8 observers in year 2). Observers were not blinded to intervention assignment, as blinding was not feasible for theater and music sessions. However, they were not involved in the collection of outcome data. Fidelity was assessed using structured checklists, and multiple rotating observers were employed to minimize bias. A summary of the reports is presented in Table 3, including interruptions, group work composition, engagement, and intervention components.

**Table 3.**
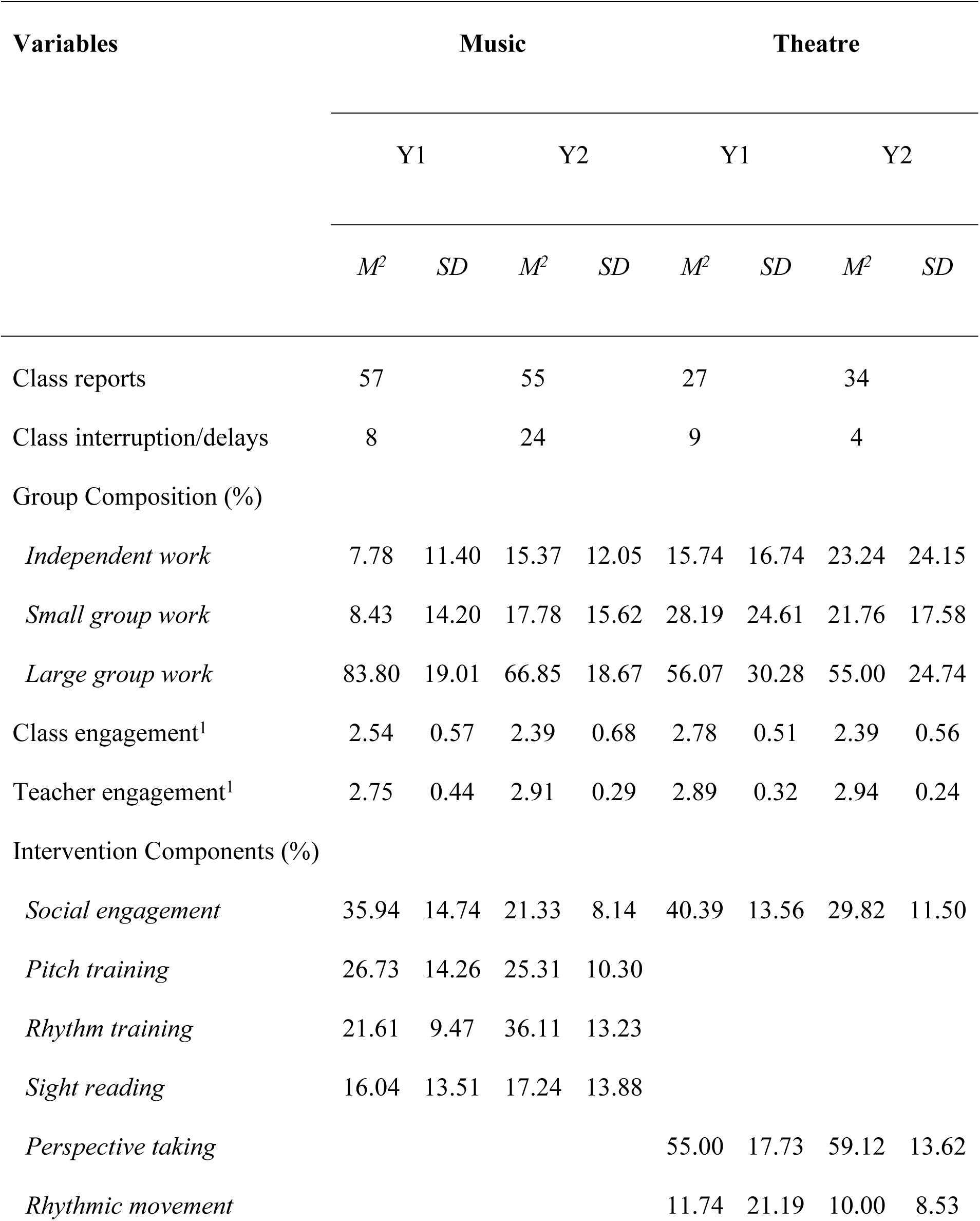
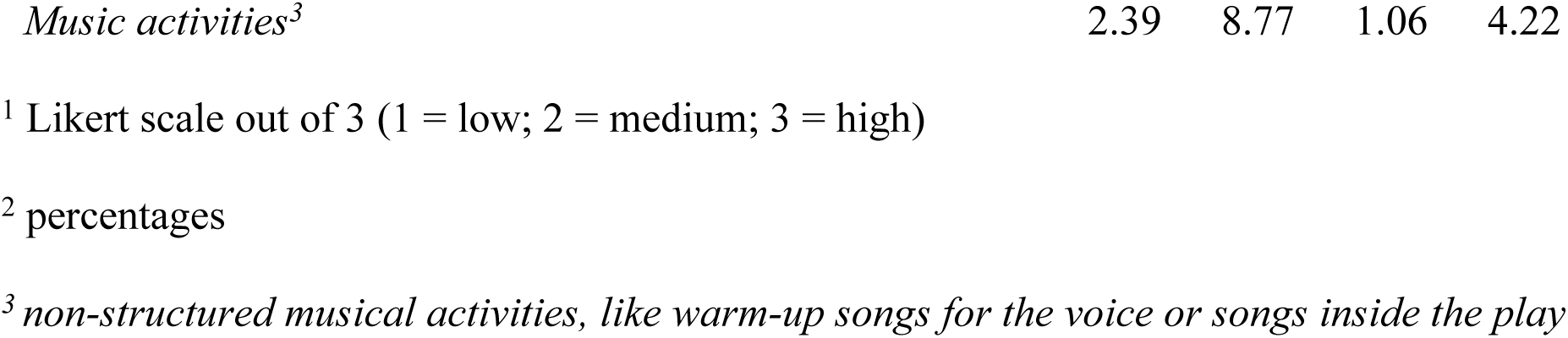
Observer reports of music and theatre intervention for year 1 and year 2 in cohort 1.

#### Cohort 2: Progress to date

To recruit cohort 2, we contacted 488 applicants about participating in the EXCEL trial. Following screening, 84 eligible and willing participants were randomized to interventions and completed baseline testing. After baseline testing, a total of 9 participants discontinued before beginning the intervention (logistics = 6; motivation = 1; other = 2). Of the 84 randomized participants, 75 enrolled in the intervention phase. At the current 6-month check-in (some are still ongoing), 55 participants (73.33%) completed follow-up assessments, and 66 are currently active on the trial in year 2, with a retention rate of 88.00%. In total, 65 of 75 active participants have asserted commitment to continue (86.67%). Among participants who completed year 1, 61 of 75 students (81.33%) met the adherence criterion, exceeding the transition threshold of 70.00%. These results indicate that the EXCEL trial met both pre-defined feasibility criteria for retention and adherence after one year of programming in the full sample.

## Discussion

This paper presented the EXCEL trial protocol and evaluated the initial feasibility of implementing an RCT of a music-based intervention for children aged 6–8, examining effects on both neural circuitry and behavioral measures of IC. Unlike prior studies that typically rely on self-selected participation in music training, the EXCEL trial uses random assignment to ensure greater experimental control and to reduce selection bias related to pre-existing interests or abilities. Demonstrating that children can adhere to and benefit from long-term training when randomly assigned is essential for establishing the intervention’s efficacy and practicality.

Participants in the EXCEL trial were randomly assigned to either a music or a theatre intervention, with the research team unaware of group allocation. The duration of the intervention was selected based on evidence that improvements in IC in children typically emerge after two or more years of training (Hennessy et al., 2019). Using an active arts-based control group, rather than a passive or no-treatment control, allows us to isolate the specific effects of music training from those associated with general group-based artistic activities. This design offers a rigorous test of music-induced changes on IC development in childhood. While this active control design provides a rigorous test of music-specific effects, such comparators may also confer cognitive or social benefits, potentially reduce between-group differences, and increase the risk of false negatives.

Analyses of feasibility outcomes (i.e., retention, adherence, and fidelity) are provided for cohort 1 and cohort 2 to help establish the viability of RCTs of music interventions and support the scalability of such approaches. Our data shows high retention (>70%) and adherence to the programs. Fidelity monitoring indicated generally high levels of implementation across both music and theatre programs, with notable variation in group composition and instructional focus. Class engagement and teacher engagement ratings remained consistently moderate to high across conditions, suggesting sustained participant interest and instructor involvement. While both interventions emphasized social engagement, the music program showed greater consistency in structured components such as pitch, rhythm, and sight-reading, as reflected in its relatively stable means and standard deviations across years. In contrast, the theatre program incorporated additional components, such as perspective-taking and, to some extent, rhythmic movement with varying intensity, as shown by the wider standard deviations, particularly in year 2. These differences reflect the distinct pedagogical aims of each program: music offers more technical training, while theatre focuses more on emotional development. Importantly, the increased number of class reports and reduced interruptions in year 2 suggest improved delivery logistics over time. Overall, these findings support the feasibility and unique characteristics of the two interventions while keeping them comparable methodologically in terms of instruction and social engagement.

An inherent limitation of longitudinal RCTs in children with an intervention such as music training is the risk that participants in the control group (theatre) will independently join a music training program. We anticipate this will be infrequent in the EXCEL trial because, unfortunately, the target population is children from families with limited access to public music education programs and private music training, which can be expensive to pursue. Moreover, the control activity (theatre) is an equally educational, creative, and motivating program that we expect to keep the students engaged and committed.

In conclusion, the EXCEL trial is a novel approach to assess the effects of music training on the development of brain IC circuitry during the critical transition from childhood to early adolescence, as well as determine the viability of longitudinal RCTs with music training in children from underserved communities. We demonstrate achievement of targets on adherence and retention after two years of intervention for an initial cohort of 42 participants. The final EXCEL trial report will include the addition of the second cohort of 75 participants (total 117) and analysis of the efficacy of long-term (2-year) music training on IC and corresponding neural changes. The EXCEL trial will form the basis for further research into this field, including the ability to expand this music programming to more children, as well as provide an example of music training as a viable and important element in child education and cognitive development.

## Data Availability and Dissemination

The datasets generated and analyzed during the current study are available from the corresponding author upon reasonable request. Planned analyses are disclosed at ClinicalTrials.gov Identifier: NCT05502939. Data will be shared to participants via media outlets and to researchers at international conferences involving music cognition and neuroscience. Data is and or will be stored on encrypted, password-protected institutional servers. All records will be de-identified prior to storage, and access will be restricted to authorized study personnel.

## Data Monitoring Committee

Given the low-risk, non-invasive nature of the interventions (music and theatre training), a formal data monitoring committee was not established. Study oversight is conducted by the principal investigators and senior research staff, who are responsible for monitoring study progress, data quality, and participant safety. The study is additionally overseen by the Institutional Review Board (IRB) at the University of Southern California.

## Author Contribution

Kevin Jamey contributed to formal analysis, writing – original draft preparation, writing – review and editing, visualization, software, and data curation. Ellen Herschel contributed to writing – original draft preparation and investigation. Caitlin Noel contributed to writing – review and editing, project administration, and investigation. Jed Villanueva contributed to writing – review and editing and investigation. Melissa Reyes contributed to writing – review and editing and project administration. Eustace Hsu contributed to writing – review and editing. Beatriz Ilari contributed to writing – review and editing, conceptualization, methodology, and validation. Wendy Mack contributed to writing – review and editing, supervision, and methodology. Shan Luo contributed to writing – review and editing, supervision, resources, methodology, funding acquisition, and conceptualization. Assal Habibi contributed to writing – review and editing, supervision, resources, methodology, funding acquisition, and conceptualization.

## Acknowledgments

The authors would like to thank the other members of our laboratory for their contributions to the success of this project. In particular, we are grateful to Cassandra Liu for her help in recruiting participants for cohort 1, Ulises Venegas-Rivera for his role in coordinating cohort 1, and Priscilla Perez and Alison Wood for their roles in data collection and general study management. To support clarity and precision in written expression, the authors used ChatGPT (OpenAI, 2025) to refine the wording of specific passages. The tool was used strictly for language enhancement, and all substantive content and interpretations remain the responsibility of the authors.

